# Comparative Effectiveness of Traditional Chinese Medicine vs. Losartan on Blood Pressure: Real-World Insights from RCT-Eligible Populations

**DOI:** 10.1101/2024.03.14.24304324

**Authors:** Kuan Jiang, Xin-Xing Lai, Shu Yang, Ying Gao, Xiao-Hua Zhou

## Abstract

When evaluating the effectiveness of a drug, a Randomized Controlled Trial (RCT) is often considered the gold standard due to its perfect randomization. While RCT assures strong internal validity, its restricted external validity poses challenges in extending treatment effects to the broader real-world population due to possible heterogeneity in covariates. In this study, we employed the augmented inverse probability of sampling weighted (AIPSW) estimator to generalize findings from an RCT comparing the efficacy of Songling Xuemaikang Capsule (SXC) —— a traditional Chinese medicine (TCM) and Losartan on hypertension reduction to a real-world trial-eligible population. Additionally, we conducted sensitivity analyses to assess the robustness of the AIPSW estimation against unmeasured confounders. The generalization results indicated that although SXC is less effective in lowering blood pressure than Losartan on week 2, week 4, and week 6, there is no statistically significant difference among the trial-eligible population at week 8, and the generalization is robust against potential unmeasured confounders.

## Introduction

Hypertension is one of the most prevalent chronic diseases globally. Losartan, a recognized medication, is often prescribed for treating hypertension^1–3^. However, in China, Songling Xuemaikang Capsule (SXC) —— a traditional Chinese herbal medicine, is recommended to treat mild hypertension clinically as an alternative in certain official guidelines^4^. Previous RCT comparing efficacy on lowering blood pressure between the two drugs demonstrated the non-inferiority of SXC to Losartan^5^. Although RCT serves as the golden standard for evaluating drug efficacy, it lacks representativeness, leading to debates when applying results to real-world situations. As evidence, there are substantive studies indicating the inconsistency between results obtained from RCT and a real-world study due to different population heterogeneity^6–9^. Compared with RCT, a registry study provides a sample more representative of the general population than RCT, but unmeasured confounders could diminish its reliability^10^. Since RCT and real-world study have their strengths and weaknesses, it is important to generalize RCT results to a real-world trial-eligible population to draw correct conclusions^11^.

There are some statistical methods available to solve the problem, which have been proven reasonable mathematically. Intuitively speaking, we can model outcomes in RCT directly and make predictions in real-world data by the fitted model (the outcome-model-based estimator)^12^ or model the sampling score (probability of a unit being sampled into RCT population) and reweight RCT results by the inverse of sampling score (IPSW)^13–15^. However, these methods need a correctly specified model to yield consistent estimation. Augmented inverse probability of sampling weighted (AIPSW) method models outcome and sampling score simultaneously and can get consistent estimation if either of the two models is correctly specified, also called doubly robust estimation^16,17^. Recently, Lee et al. proposed a calibration weighting method to reweight units in the RCT sample,and after calibration, the covariate distribution of the RCT sample empirically matches the real-world trial-eligible population^18^. These methods generalize RCT results to a real-world trial-eligible population by the following data structure: treatment assignment in RCT, the RCT outcomes, and observed common covariates in both RCT and real-world data. For reviews on existing methods, readers can refer to them for formulas and relative proofs^17,19^.

In addition to addressing observed covariates imbalance between RCT and real-world trial-eligible populations, exploring unobserved covariates is crucial and challenging for ensuring reliable generalization results by corresponding sensitivity analysis. Nguyen et al. proposed a set of methods to evaluate the bias of IPSW and model-based estimators when unobserved confounders exist. Still, the methods rely on a strict parametric setting^20^. Recently, based on Cinelli et al. and Douglas et al., Huang proposed several tools to assess the robustness of generalization results, including numeric statistics (robustness value, minimum relative confounder strength, etc.), bias counterplots, and a formal benchmarking approach. The advantage of Huang’s method is that it does not require any assumptions on the data-generating process^21–23^.

In this article, we generalized the results in RCT comparing SXC with Losartan on the efficacy of treating hypertension in the real-world trial-eligible population through the AIPSW estimator. We found that conclusions in RCT and real-world populations were different. In RCT, there was no statistically significant difference in lowering systolic blood pressure (SBP) or diastolic blood pressure (DBP) between the SXC and Losartan group at all follow-up visits. However, in the real-world trial-eligible population, Losartan was more effective in lowering SBP in the early stages (weeks 2, 4, 6) but had a non-superior effect to SXC on lowering SBP and DBP at week 8. To address potential threats from unmeasured confounders, we performed a sensitivity analysis to assess the robustness of non-inferiority at week 8 by tools proposed in Huang et al. Corresponding sensitivity analysis on week 8 estimation indicated the robustness of generalization results against potential violation of assumptions. Therefore, SXC was not inferior to Losartan in lowering blood pressure (BP) in real-world trial-eligible populations in the long term.

## Method

### Data Source

We conducted two studies: an RCT and a registry study. The RCT was a multicenter, randomized, double-blind, non-inferiority trial that compared SXC with Losartan in terms of the efficacy of lowering blood pressure. It enrolled patients 18 to 65 years of age with mild essential hypertension (Grade I hypertension). In the treatment group, patients were assigned to receive SXC monotonously, and patients in the control group were assigned to receive Losartan. Details of the study design and the primary results were published previously^5^. The registry study collected patients in the real-world practice setting who used SXC monotonously or a combination of SXC and other medicines to lower hypertension. We gathered baseline covariates and conducted follow-up visits at weeks 2, 4, 6, and 8 in both studies. The study protocol was approved by the Institutional Review Board of Dongzhimen Hospital affiliated to Beijing University of Chinese Medicine (approval number: ECSL-BDY-2011-19) and was registered on the Chinese Clinical Trial Registry Platform (www.chictr.org.cn; Unique identifier: ChiCTRONC-11001612)

### Study Objective

The primary objective of the study is to combine the RCT with real-world registry datasets to generalize RCT comparison results to a real-world trial-eligible population through the AIPSW estimator. Moreover, since the assumptions of generalization methods can be violated by unmeasured covariates, the robustness of the inference was assessed through corresponding sensitivity analysis.

### Statistical Analysis

#### Common Variables Extraction, Missing Value Imputation and Trimming

In the data preprocessing stage, we first extracted common baseline variables in RCT and real-world datasets to make the two datasets comparable. Because there are abnormal and missing values in variables ‘weight’ and ‘marriage’, we first deleted the abnormal values, treated them as missing values, and imputed them by multiple imputation. Because variables with missing values are continuous and binary, we chose the random forest method to impute the value due to its superior handling capabilities. Concerning the inclusion/exclusion criteria in RCT, we respectfully declined to include patients who didn’t meet them in the real-world study.

### Outcome

In the RCT, the outcome was defined as an effect on lowering blood pressure, calculated by subtracting baseline systolic/diastolic blood pressure (BSBP/BDBP) from observed blood pressures at 2,4,6,8-week follow-up visits. Efficacy differences between treatment and control groups were defined as the differences in means between the two groups. If the difference is greater than zero, then the effectiveness of SXC on lowering BP was worse than that of Losartan.

### Generalization of RCT results in the real-world population

We built two logistic regression models to estimate the weights: one is to regress the sampling variable (binary variable, 1 if in RCT dataset, 0 otherwise) on covariates in the combined dataset, and the other is to regress treatment assign variable (binary variable, 1 if in SXC group, 0 Losartan group) on covariates in the RCT dataset. We use the random forest to model the potential outcomes of two groups in the RCT dataset. Then we calculated the point estimation of RCT generalization results using the AIPSW estimator. Details about the formula can be found in section 5.3 in Dahabreh et al.^17^. Meanwhile, as a statistical method, the estimator depends on some necessary assumptions to get correct generalization results, which are based on potential outcome framework in causal inference^24^. Some of the assumptions (Consistency assumption, Mean exchangeability assumption in RCT, Positivity of treatment probability in the trial, Positivity of trial participation probability) make sense obviously in our setting, while a crucial assumption——the mean generalizability assumption is hard to check. Further illustrations of the assumptions are relegated in Appendix S2.

To estimate the standard error and 95% confidence interval (CI) of the above estimators, we used the bootstrap method, which repeats resampling 1000 times. If 95% CI contains 0, then it is not statistically significant at 0.05. Statistical analysis was performed using R version 4.2.2 for Windows.

### Sensitivity analysis

To ensure robust estimation against unmeasured variables that may violate the mean generalizability assumption, we conducted a sensitivity analysis on the AIPSW estimator. We assessed the robustness in the following aspects. First, we used robustness value (RV) to measure the strength a confounder needs to cause the bias equal to estimation using observed variables. Then, we examined the plausibility for such an unmeasured variable to exist by using observed covariates as a benchmark. We used MRCS, 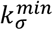 and 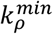 to evaluate the relative strength a potential confounder needs to disturb the estimation. Intuitively, MRCS measures how much the relative confounding strength an omitted variable must have to result in a killer confounder, and 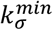 and 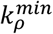 can be similarly interpreted. We described the benchmarking process on a bias contour plot, where the benchmark variables were annotated by their confounder strength. For simplicity, we relegated the detailed descriptions of the above measurements to Appendix S2, readers can refer to them for further explanation.

## Results

### Descriptive Analysis

We initially recruited 602 patients with grade I hypertension into the RCT, 300 of whom were assigned to the treatment group randomly. Meanwhile, 3,000 patients were enrolled in a registry study, and they only used SXC (N = 1567) or a mixture of SXC and other Western medicine (N = 1433) to lower blood pressure. After trimming the real-world data, 804 patients were left in the real-world cohort as a trial-eligible population.

### Baseline covariates at visit

We extracted seven common variables in both datasets: Age, Sex, Body Mass Index (BMI), Marriage, Smoking, BSBP, and BDBP. Summary statistics of the variables are presented in Table 1. Of the baseline covariates, the mean of age was significantly greater in the real-world dataset (RWD) population than in the RCT group but was balanced between the treatment group and the control group within the RCT population. The standard deviation of BDBP in the RWD population is greater than the RCT population. Other baseline covariates were similarly distributed. Kolmogorov-Smirnov test (K-S test) was performed on continuous variables, and p-values were less than 0.05, indicating that the distribution of variables between the two populations was significantly different at the 0.05 level. Similarly, binary variables were tested by a two-sample Z-test. Box plots of continuous covariates were provided in Figure 2. In the real world, people using SXC were older than those in RCT and had lower BDBP than in the real-world trial-eligible population.

**Table 1.**
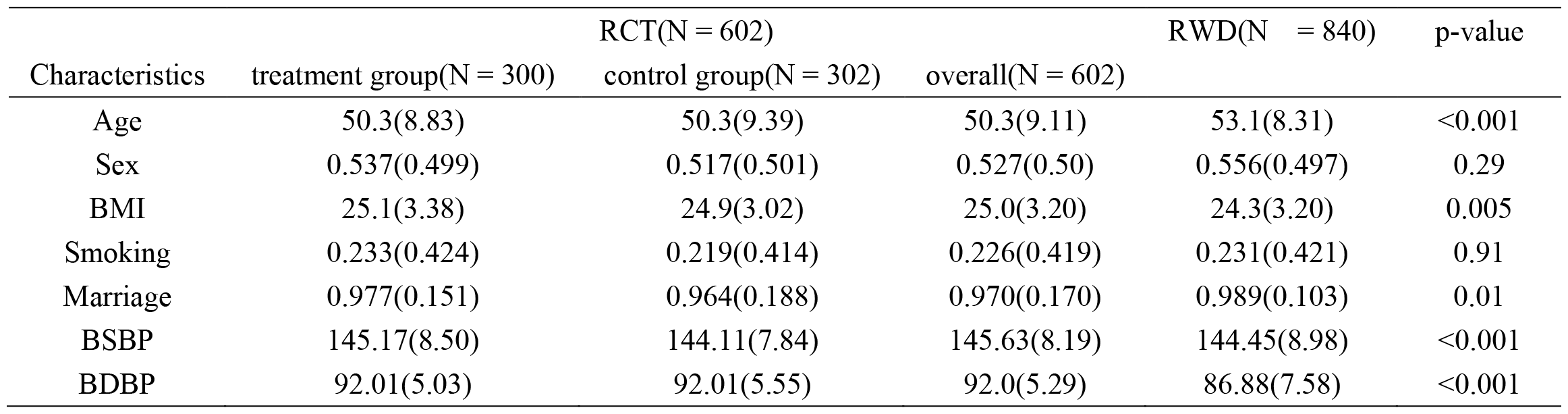
characteristic of baseline covariates and test between RCT and real-world data (RWD) population.

**Fig 1.**
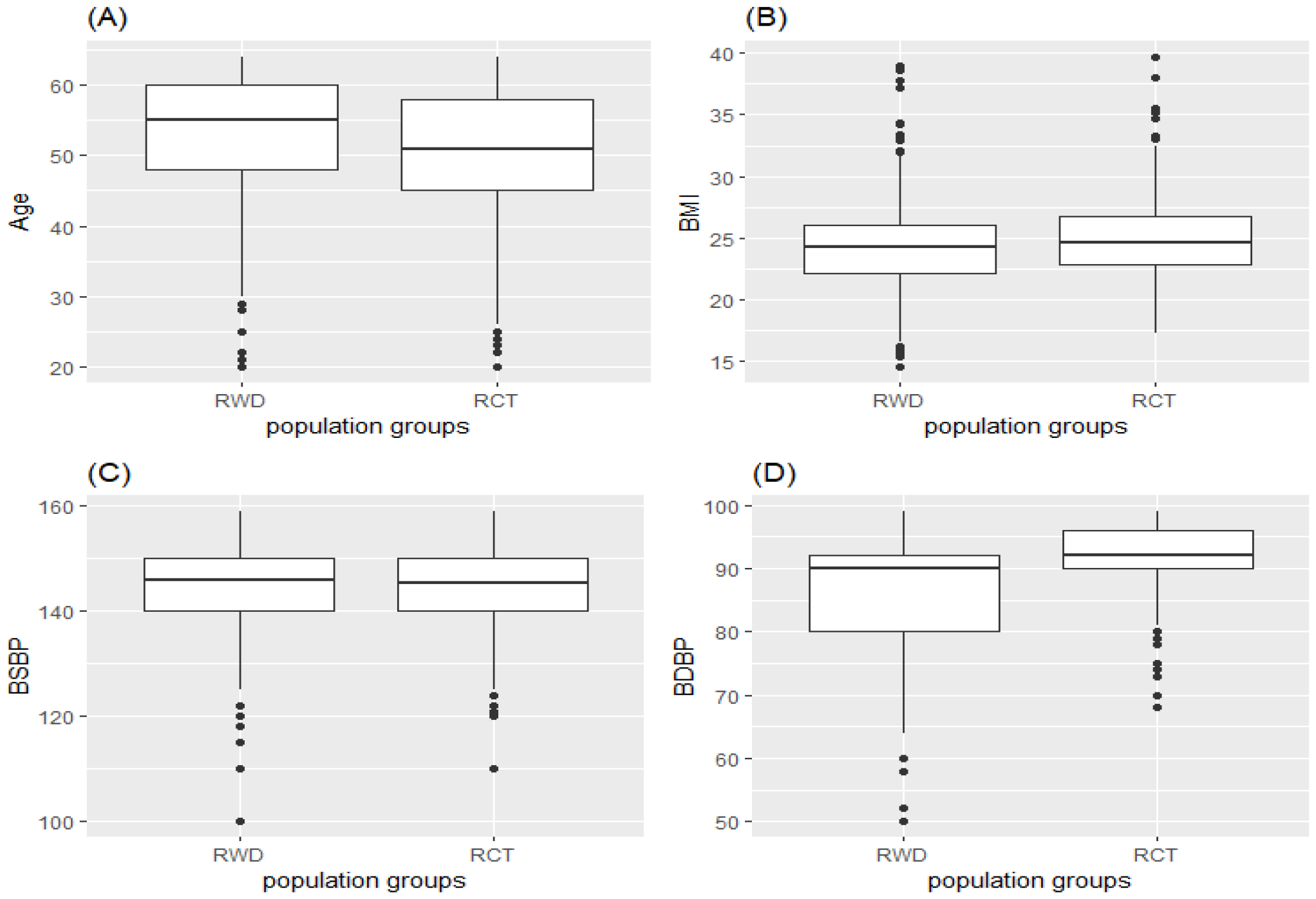
Box plots of continuous baseline variables in RWD and RCT sample: (A).Age, (B).BMI, (C). BSBP, and (D).BDBP

**Fig 2.**
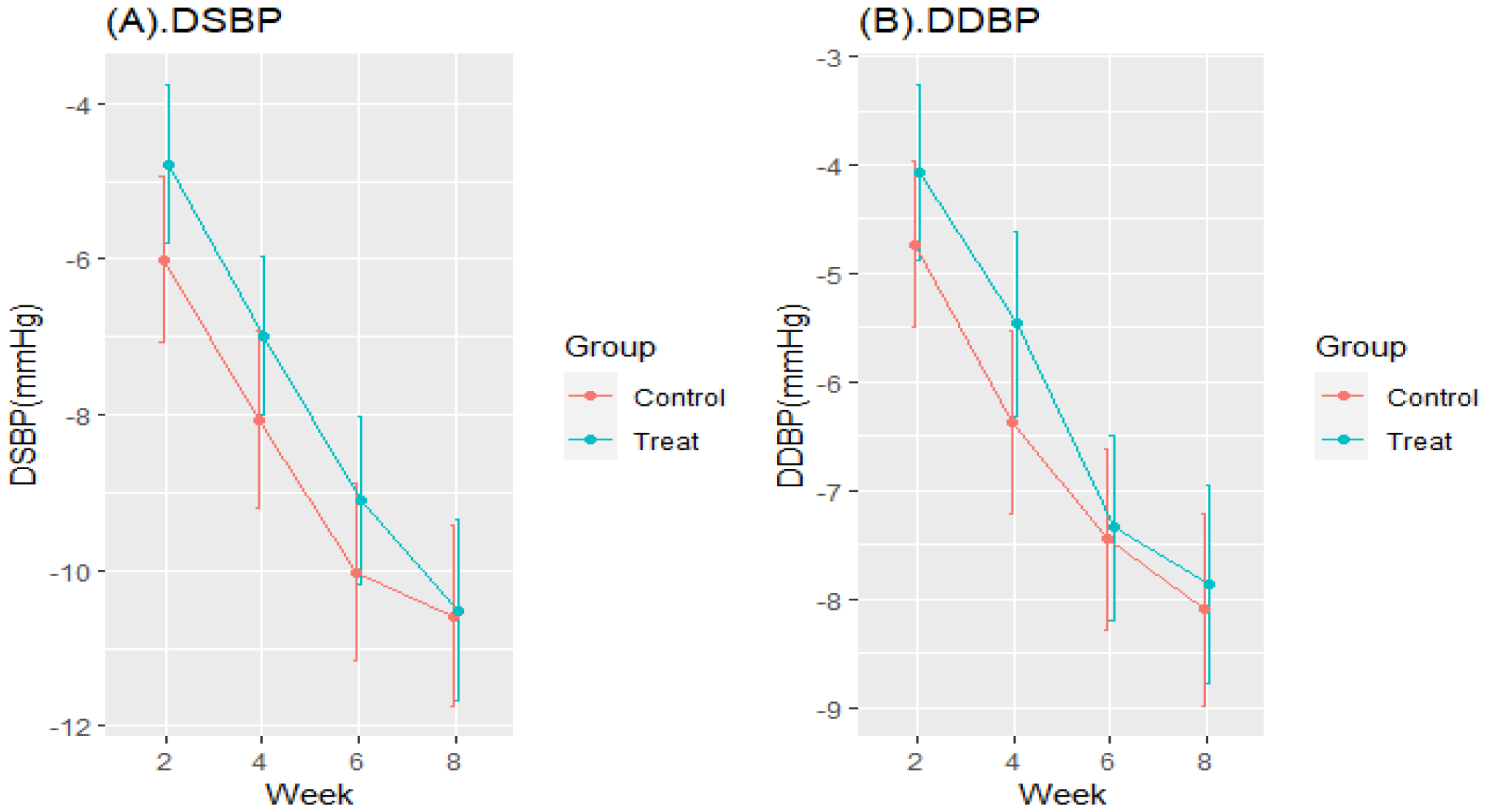
Difference of blood pressure at following-up visits in RCT population. (A). DSBP (B).DDBP

### RCT results

We presented the difference between SBP at follow-up visits and BSBP (DSBP) in the treatment and the control group in Figure 2(A) and DDBP in Figure 2(B) similarly. Compared to Losartan (control group), SXC was less effective in lowering SBP/DBP in the short term (weeks 2, 4, 6) but had a similar effect in the long term (week 8). The difference between the effect and 95% CI was provided in Table 2, and all CIs contain 0, implying that the difference between the two groups was not statistically significant at 0.05. Thus, we concluded that SXC was not inferior to Losartan in curing hypertension in the RCT population.

**Table 2.** Difference of efficacy in lowering BP between the treatment (SXC) and the control group (Losartan) in RCT.

| Week | DSBP |  | DDBP |  |
| --- | --- | --- | --- | --- |
|  | point estimation | 95% CI | point estimation | 95% CI |
| 2w | 1.230(0.811) | (-0.370,2.822) | 0.662(0.593) | (-0.591,1.736) |
| 4w | 1.080(0.773) | (-0.400,2.627) | 0.911(0.644) | (-0.318,2.079) |
| 6w | 0.920(0.781) | (-0.537,2.466 ) | 0.104(0.643) | (-1.251,1.358) |
| 8w | 0.073(0.858) | (-1.674,1.759) | 0.239(0.670) | (-1.103,1.394) |

### RCT generalization to RWD

Comparison of the two drugs on lowering SBP and DBP in RCT and its generalization to real-world trial-eligible populations are in Figures 3(A) and 3(B). On lowering SBP, although no statistically significant result was reported in RCT when generalized to the real world, the difference was significant except at week 8. In contrast, RCT and AIPSW estimations were similar in lowering DBP except at week 2, where AIPSW estimation was slightly significant. The comparison showed a discrepancy in conclusion between the two populations. In RCT, the efficacy of SXC on lowering SBP was comparable to Losartan, while in real-world trial-eligible populations, the conclusion held only in the long term (8w). As for DBP, conclusions from RCT and its generalization were similar, where SXC was not inferior to Losartan.

**Fig 3.**
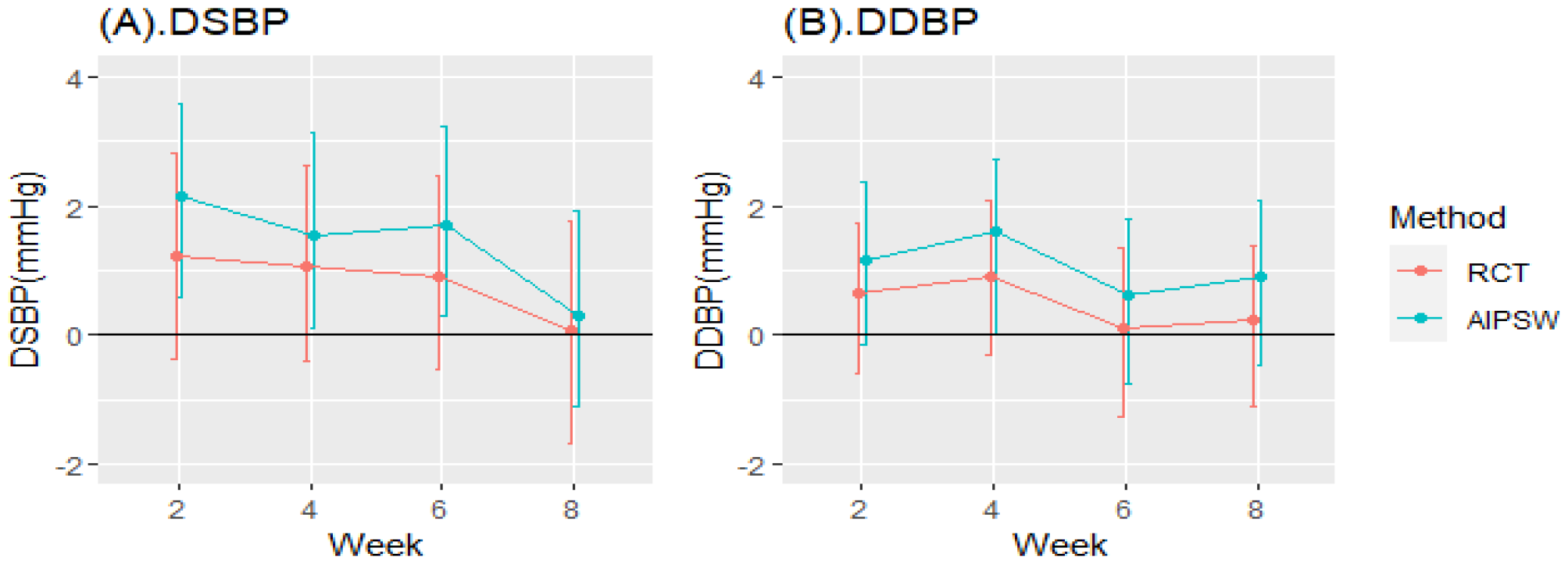
Difference of efficacy on lowering BP in RCT and its generalization to the real-world trial-eligible population by AIPSW estimator

### Sensitivity analysis

We performed sensitivity analysis on the upper and lower bounds of the AIPSW estimation in week 8. Table 3 reported the minimum relative strength over benchmark variables for an unmeasured variable to be a killer confounder for the lower bound, which means that the confounder can overturn the negative value to zero or even positive if observed. Once the lower bound becomes positive, we can conclude SXC’s significant inferiority to Losartan in lowering blood pressure, then the generalization result on week 8 will be unreliable. Take DSBP as an example; the required relative strength (i.e. the ratio of bias caused by omitted confounders over benchmarking variables) for an unobserved confounder is no less than 58.16 times gender, 24.77 times age, and 23.2 times marriage, etc., to overturn the negative lower bound. Since we had collected all key confounders according to our clinical practice, we were confident in denying the existence of such variables. A similar conclusion was for DDBP at week 8.

**Table 3.**
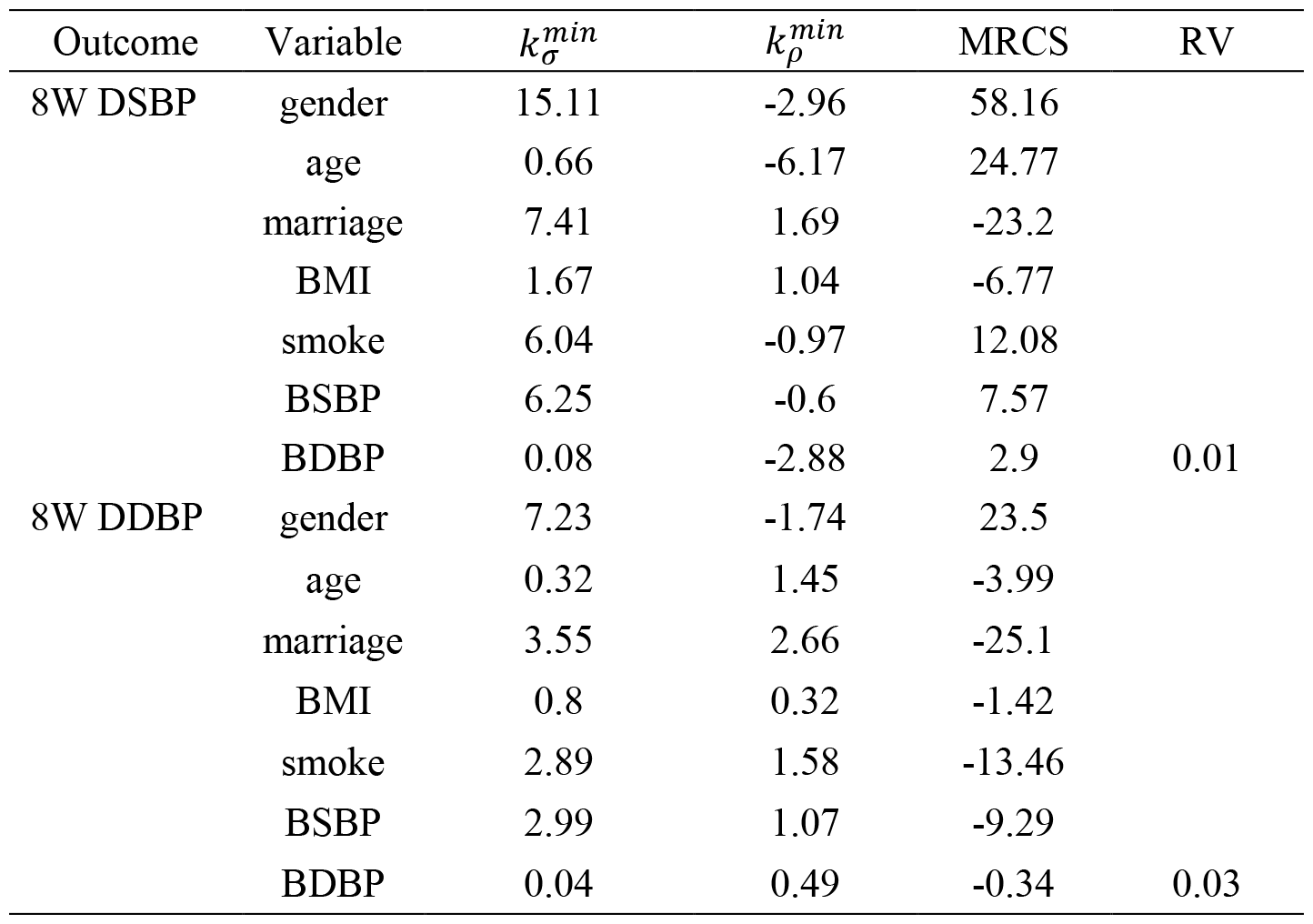
Summary statistics and relative strength to benchmarking variables for AIPSW lower bounds at week 8.

The bias contour plot for the AIPSW estimation bounds and confounding strength of benchmarking variables for the lower bound at week 8 were provided in Figure 4(A) and Figure 4(B). In Figure 4(A), if unmeasured confounders exist in the green zone, then the upper bound can be reversed, indicating SXC’s superiority over Losartan. In contrast, the lower bound can be reversed by unmeasured confounders in the red area, meaning that SXC is inferior to Losartan. In Figure 4, all the benchmark variables were not in the red area, and we concluded that our estimation was robust against potentially unmeasured confounders. This was reasonable because when designing clinical trials, we attempted to gather all major confounding variables and believed there were no stronger confounders than what we have included based on clinical practices.

**Fig 4.**
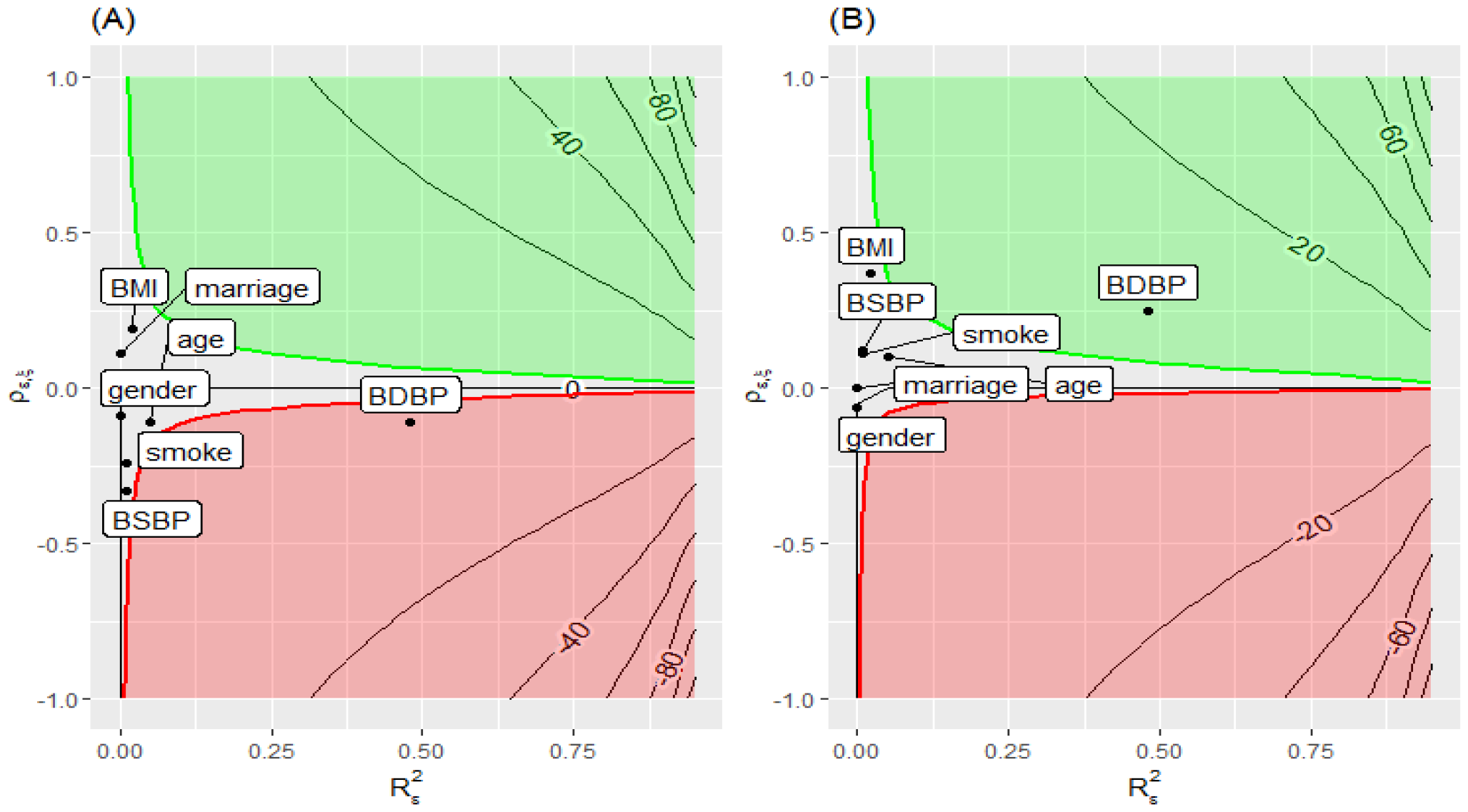
Bias plots for AIPSW bounds at week 8. Benchmarking variables are for the lower bound.

## Discussion

In this article, we compared the efficacy of lowering blood pressure using SXC and Losartan in RCT. We used the AIPSW estimator to generalize the results to a real-world trial-eligible population. We found inconsistency between the RCT and its generalization results. Although the difference between the two drugs in treating hypertension was statistically insignificant in RCT, the efficacy of lowering SBP at week 2, week 4, and week 6 of SXC was inferior to Losartan significantly when it was generalized to the real-world trial-eligible population. As for lowering diastolic blood pressure, SXC was inferior to Losartan at week 4, but similar at week 6 and week 8. In the long term, we can conclude that SXC is not inferior to Losartan in treating hypertension in the real-world trial-eligible population. The difference in SXC efficacy in two populations can be attributed to covariate distribution shifts. Take age as an example. From Figure 1 and Table 1, the real-world population has more older patients than the RCT population. This may explain the difference in the study conclusions to some extent, perhaps because SXC works to lower blood pressure for seniors in the long term compared to Losartan. However, there was a higher proportion of older patients in the real-world trial-eligible population, thus SXC behaved less effectively than RCT in the short term. In terms of generalization results, our analysis generated important hypotheses for further exploring the mechanisms of how SXC works and served as a guideline for using SXC to treat hypertension in the real-world population. In the real world, using a mixture of SXC and Losartan to treat hypertension may be a better solution to balance efficacy and side effects, which had been proven in existing studies^25^.

The discrepancy between RCT and the real-world study was mainly due to covariates heterogeneity between RCT and real-world populations, including those that cannot be measured or observed directly. This has led to the inability of RCT populations to accurately represent trial-eligible populations in real-world contexts. There are numerous studies revealing the heterogeneity, many of which used matching methods to compare results obtained from RCT and real-world studies^6–8^. For instance, a previous study utilized the propensity score matching (PSM) method to match SXC users in both RCT and real-world cohorts and concluded the SXC’s non-inferiority in treating hypertension in real-world population to RCT population. In their framework, after balancing observed outcomes, the heterogeneity comes from external unmeasured covariates. However, to generalize RCT results to the real world, the imbalance between the observed variables of the RCT population and the real-world trial-eligible population must be considered. Therefore, in our study, we sought to address heterogeneity from both observed and unobserved variables simultaneously. We first used the AIPSW estimator to make a generalization, which addressed the covariates distribution shift between the two populations under study. However, the statistical method depends on the mean generalizability assumption, which is potentially threatened by unobserved variables. Therefore, we made a sensitivity analysis to illustrate that our generalization results were robust against heterogeneity from unobserved variables.

In addition to the AIPSW estimator, we applied other methods introduced in the previous section to generalize the RCT results. The corresponding estimation tables and figures can be found in supplemental materials. As shown in Figures S3.1 and S3.2, despite the similarity of point estimation, the three methods exhibited different standard errors. The difference in standard errors was explained by Dahabreh et.al., that when the probability model and outcome model are correctly specified, the large-sample variance of AIPSW estimators will be larger or equal to that of outcome model-based method and no larger than that of IPSW estimators^17^. The explanation was in accordance with our practical results.

There are still some limitations in our study. Firstly, we relied on statistical generalization rather than evidence-based generalization, meaning we could not access real-world trial-eligible population data to verify our findings. Obtaining such evidence would be challenging, as designing RCT on the target population would be difficult, and conducting real-world studies may be affected by unmeasured confounders, which could introduce bias. However, the mathematical property of the estimator ensured that our results were reliable even in the absence of real-world evidence. Secondly, in the study, we dropped outpatients in the real-world cohort who did not meet the inclusion/exclusion criteria of the RCT to make a more accurate estimation. Therefore, our generalization conclusion was limited to the trial-eligible population. Recently, Paul et al. proposed a synthesis parametric model to deal with the non-positivity generalization problem, but this method requires subjective information, making it challenging to apply in the practice field^26,27^.

## Conclusion

We concluded that although in the RCT population, the SXC was non-inferior to Losartan in lowering SBP and DBP, in real-world trial-eligible population, the SXC is inferior to Losartan in lowering SBP in the short term but non-inferior in the long term. Besides, our conclusion in week 8 (the non-inferiority conclusion) was robust, against potentially unmeasured confounders.

## Data Availability

The author cannot share the data without a permit from a relative institution.

## Acknowledgments

We thank all patients and their families for participating in the registry or the SXC-BP trial and all the centers and investigators who devoted their effort and time to the two studies.

